# How does news coverage of suicide affect suicidal behaviour at a high-frequency location? A seven-year time-series analysis

**DOI:** 10.1101/2024.10.02.24314762

**Authors:** Lisa Marzano, Ruth Spence, Ian Marsh, Arianna Barbin, Ian Kruger

## Abstract

**Introduction:** News reporting of suicide can have a significant influence on suicidal behaviour in the general population, especially following the death of a well-known individual. By comparison, the impact of reporting on suicides at well-known, ‘high-frequency’ locations are less well understood. We investigated the relationship between news coverage of suicide and incidents at a high-frequency coastal location in the UK over a seven-year period.

**Methods:** We analysed bi-directional associations (with daily and weekly lags) and Granger causality between suicide-related news in the UK (N=38,595, of which 789 focusing on cliff locations) and suspected suicides (N=278) and crisis interventions (N=3,050) at the site between 1^st^ January 2017 and 31^st^ December 2023. Separate sub-analyses explored associations with repeat coverage and with headlines featuring explicit location/method details.

**Results:** Whilst coverage of incidents at the study site and other coastal locations represents a small and decreasing proportion of all UK news of suicide, 51% of all cliff-related news focused on the study site, often explicitly identified in the story’s headline (81%). There were significant but small correlations between volume of news coverage (particularly when method– and location-specific) and suicidal behaviour at the site, with fatalities increasing in the immediate aftermath of reporting. This effect was strongest in 2018-19 (which had the greatest volume of reporting and repeat coverage), but failed to reach significance in 2020-2023, when there were fewer reports, less repeat coverage, and no headlines referring to multiple deaths at the site.

**Conclusions:** Our findings underscore the importance of continued efforts to monitor and improve the quality of news and other media portrayals of suicide. Follow-up studies, including qualitative research with people with lived/living experiences of suicide, could further explore how different types of news stories and wider narratives might contribute to increases – and potentially decreases – in suicides at high-frequency locations.

**Key Messages:** *What is already known on this topic:*

○ Associations between news coverage of suicide and increases in suicidal behaviour have been well documented. However, recent evidence suggests that their relationship may be more complex than originally thought, and limited to specific types of reports.
○ A recent meta-analysis found that reporting of celebrity suicides can have a meaningful influence on suicides in the general population. In contrast, general reporting of suicides appears not to be associated with suicide – but fewer studies have investigated their impact.
○ Media portrayals that describe suicide methods have been associated with increases in suicides, but the impact of reports focusing on suicide locations – including ‘high-frequency’ locations – is less well understood.
○ We therefore investigated associations between general reporting of suicide, as well as method– and location-specific news coverage, on fatal and non-fatal suicidal behaviour at a high-frequency location in the UK.

*What this study adds:*

○ We identified small but significant associations between news coverage of suicide (particularly when method– and location-specific) and suicidal behaviour at the study site. This includes increases in suspected suicides in the immediate aftermath of reporting (generally within a day or two, and no more than 9 days later), and in crisis interventions in response to increased general reporting (regardless of method/location details).
○ Our findings also point to the importance of volume of reporting and repeat coverage of ‘high-impact’ stories in assessing the potential impact of news coverage.

*How this study might affect research, practice or policy:*

○ Guidelines on responsible reporting of suicides are an important component of national suicide prevention strategies worldwide. It is therefore crucial that these reflect high quality, up-to-date evidence, relating to a range of reporting.
○ Our findings have implications for reporting on suicides at high-risk locations, and follow-up studies to better understand the influence of different story-types and wider narratives on different audiences.

## Introduction

Research has repeatedly shown that media reporting of suicide can influence suicidal behaviour and lead to imitative acts, particularly when the coverage is extensive and sensationalist, and when overt description of the method(s) of suicide is included [1].

The association between suicide reporting in the media and suicide appears to be particularly strong following coverage of a celebrity suicide, especially when the suicide method used by the celebrity is reported [2]. By comparison, the impact of reports focusing on well-known locations (as opposed to well-known individuals) has received less attention in the literature. An important exception is an Austrian study (and follow-up research) which highlighted significant decreases in suicides and attempts on the Vienna underground railway system following the introduction of guidance on responsible reporting of such deaths [3–5].

Current international media guidelines explicitly warn against “naming or providing details of the site/location [of a suicide]” [6], as this is feared to affect the number of subsequent suicides at the same location. As recommended by the Samaritans’ media guidelines for reporting suicide, “don’t refer to a specific site or location as popular or known for suicides, for example, ‘notorious site’ or ‘hot spot’ and refrain from providing information, such as the height of a bridge or cliff” [7].

These and equivalent guidelines in other countries do not, however, suggest a blanket ban on media coverage of suicide. A growing body of evidence suggests that media reports can (also) have beneficial effects, when focused on stories of hope and recovery [8]. In other words, “media reporting of suicide can lead to subsequent increases in suicidal behaviours *if* the reporting is not consistent with best practices” [6; emphasis added].

The current study aimed to further understanding of the relationship between news reports and suicidal behaviour at the location with the highest frequency of suicides in the UK. Using suicide news data and information about suspected suicides and crisis interventions at this coastal site, we explored daily and weekly associations over a seven-year period. In addition to analyses based on the volume of reporting, we investigated associations with coastal suicide news stories featuring explicit method and/or location details in the headline, and following extensive coverage of single, high-profile incidents. Both types of reporting are actively discouraged in national and international media guidelines [6,7] and were therefore hypothesised to have a greater impact on suicides and life-saving interventions at the location.

## Materials and Methods

### Data Sources

Our dataset comprised daily time series data from 1^st^ January 2017 and 31^st^ December 2023, pertaining to:

#### 1. News of Suicides and Suicide Attempts in the UK Press (both Print and Online)

To extract this information, we used the Samaritans’ media monitoring database, which includes details of online and press newspaper articles about suicide and attempted suicide in the UK, coded for identifying information (e.g. the name of the specific newspaper and article title), descriptive information (e.g. the date of the article; details on its content and genre, including method of suicide), as well as quality ratings for each item, using a set of dimensions that operationalise criteria in media guidelines (e.g. whether the article includes excessive detail of the method or methods used) [9].

For the current study, we carried out some additional coding of this database, to identify all news stories relating to cliff suicides, and to specific coastal locations in the UK and internationally. We also recoded the headlines of all coastal suicide news to identify those which included explicit method and/or location details, and examined patterns of repeat coverage in relation to the study location, based on both weekly volume of stories and repeat coverage of specific incidents.

#### 2. Suicidal Incidents at the Study Site

We were granted access to anonymised information about cliff-related incidents at the study site, by date of incident. This included data on suspected suicides (as recorded in the County’s Real-Time Surveillance System [10]) as well as information about crisis interventions (as recorded by an established search and rescue charity which regularly patrols the site and responds to emergency calls locating anyone at risk).

## Data Analysis

Following initial descriptive analyses (e.g. to identify the yearly number and percentage of news items on 1) all suicidal behaviour, 2) all cliff suicides, and 3) suicidal behaviour at the study site and 4) immediately neighbouring cliffs, we conducted a series of correlations (with daily and weekly lags) to explore the relationship between news coverage of suicide and a) suspected suicides at the study site and immediately adjacent cliffs and b) crisis interventions at the study site.

We conducted tests of seasonality and autocorrelation of the data, to ensure the time series data were stationary and suitable for using Granger causality tests [11].

Finally, we used Granger causality to assess whether temporal changes in suspected suicides, interventions and media reporting predicted subsequent temporal changes in each other. Granger causality is a statistical hypothesis test to determine whether time series *Y* can be better predicted using the histories of both time series *X* and *Y* than it can by using the history of *Y* alone (i.e. whether time series *X* ‘Granger-causes’ *Y*). The analysis gives a *p*-value and a lag value, which is the number of time intervals of a specified unit (e.g., days, weeks) after which changes in *X* are seen in *Y*. A 0.05 significance level (i.e., p<0.05) was adopted in all statistical tests.

## Patient and Public Involvement

Patients and the public were not involved in the design, conduct or dissemination of the study.

## Results

### Descriptive Analysis of Yearly News and Incident Data

Table 1 below provides a yearly breakdown of the number of suspected suicides and interventions at the study site and immediately adjacent cliffs, along with the volume of UK news of suicidal behaviour (by all methods; at coastal locations; and specifically at the study site and surrounding cliffs).

**Table 1.**
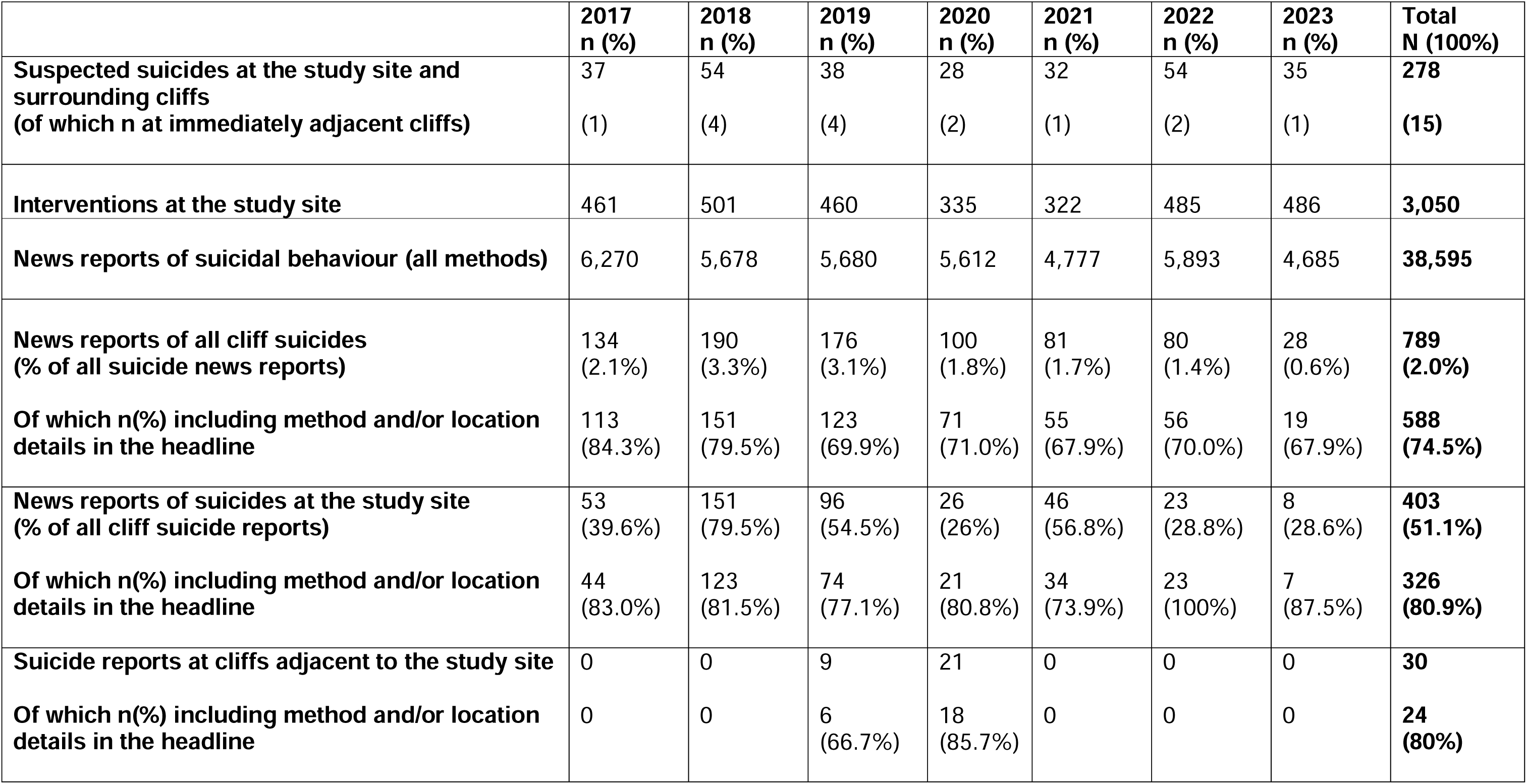
News reports of suicidal behaviour and suspected suicides and interventions at the study site, by year.

Whilst reports of suicides at coastal locations account for a relatively small proportion of all UK news relating to suicide (ranging from 3.3% in 2018 to 0.6% in 2023), it is notable that just over half of these stories focused specifically on the study site (with yearly variations).

Of note is also the substantial decrease in the number and percentage of news reports of suicides at cliff locations, and especially the study site, over the study period – despite higher-than-average suspected suicides (in 2022) and interventions (in 2022 and 2023).

However, the proportion of stories featuring location and/or method details in the headlines remained considerably high throughout. 192 of the 403 news stories focusing on the study site (47.6%) featured the cliff’s name in the headline itself; 72 made explicit reference to a nearby town; 14 named nearby specific locations within the cliff itself, whilst a smaller number of reports referred to incidents at a ‘notorious suicide (n=6) or beauty (n=4) spot’.

### Reporting of Suicidal Behaviour at the Study Site

Most reports relating to the study site featured online (327/403, 81% vs. 76,19% in print news), in local and regional press (267/403, 66.3%, of which almost half (129/267, 48.3%) from a single publication), followed by tabloid news (109/403, 27%). Approximately half the stories were reported at the time of the incident (198/403, 49.1%; vs. inquest (159, 39.5%) or other (46, 11.4%)), and nearly all were reports of fatalities (389, 96.5% vs. 14 (3.5%) stories of non-fatal suicidal behaviour).

Almost a quarter of all stories (91, 22.6%) focused on one of two ‘murder-suicides’ which took place in 2018; 26 were reports of multiple (but apparently unrelated) suicides; 1 focused on a suspected suicide pact in 2020 (later found to have been deaths by misadventure); and the remaining 285 (70.7%) on individual deaths (271) or attempts (14).

### Suspected Suicides and Interventions at the Study Site

Our analysis suggests small but significant correlations between suicides and interventions at the study site, indicating that an above-average number of suicides and immediately adjacent cliffs is significantly associated with an above-average value of interventions at the same time (r=0.33), a month later (r=0.38) or two months later (r=0.36).

**Figure 1.**
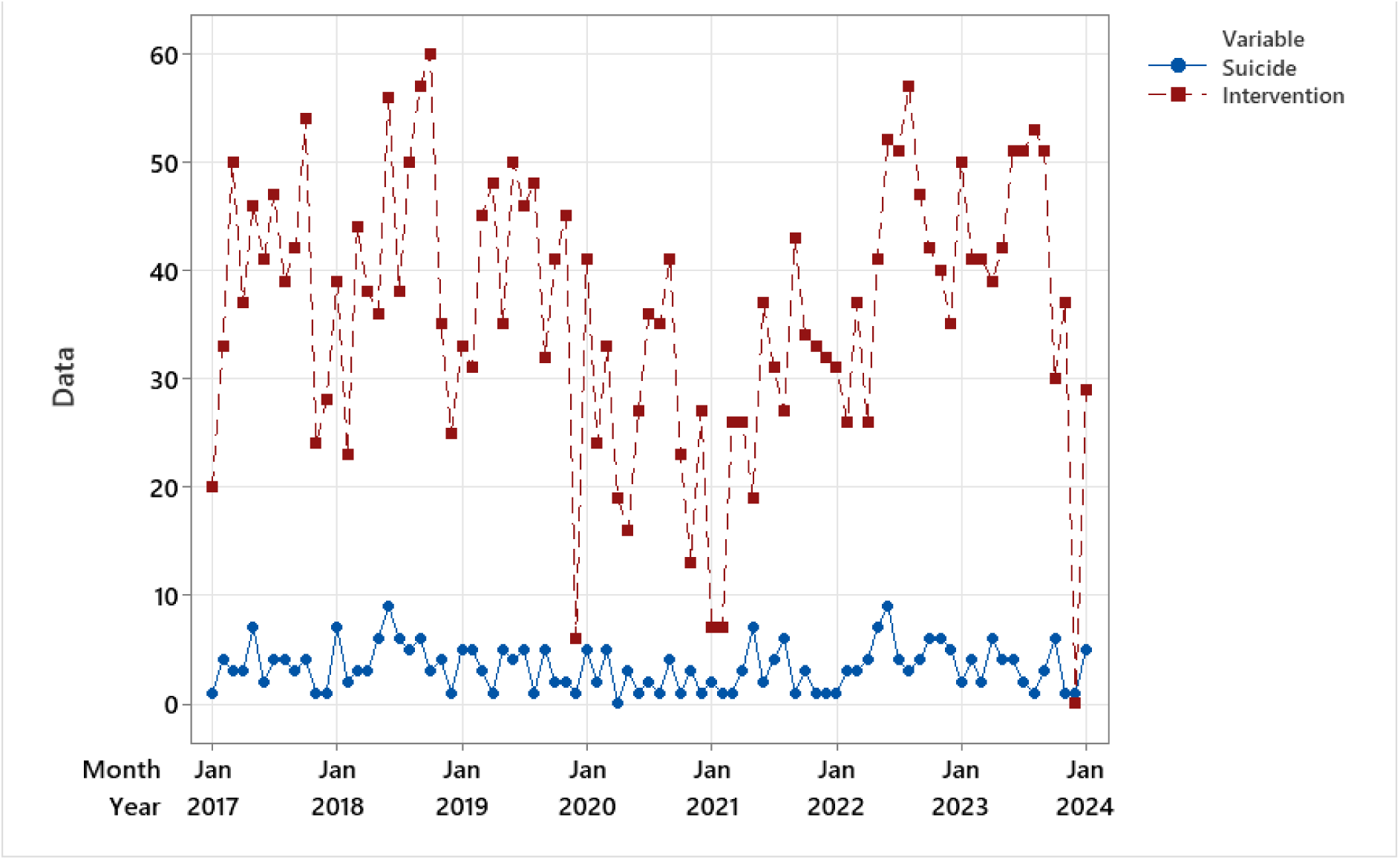
Time series plot of suspected suicides at the study sites and immediately adjacent cliffs and interventions at the site.

A Granger causality test with a month lag suggests that knowing the number of suicides is useful for predicting the future number of interventions (F=7.23, p=0.008). However, knowing the number of interventions did not appear to be useful for predicting the future number of suicides (F=0.51, p=0.48).

The number of suicides at the study site was also not significantly predicted by earlier suicides at the location, and we found no evidence of ‘seasonality’ in the data, despite some evidence of there being more suicides in May and June.

### Relationship between Media Reports and Suicidal Behaviour at the Study Site

Figure 2 below displays monthly counts of suicide news (by all methods), cliff suicide news (including specifically of suicides and attempts at the study site) and the volume of interventions and suspected suicides at the study site and immediately adjacent cliffs, between 1^st^ January 2017 and 31^st^ December 2023.

**Figure 2.**
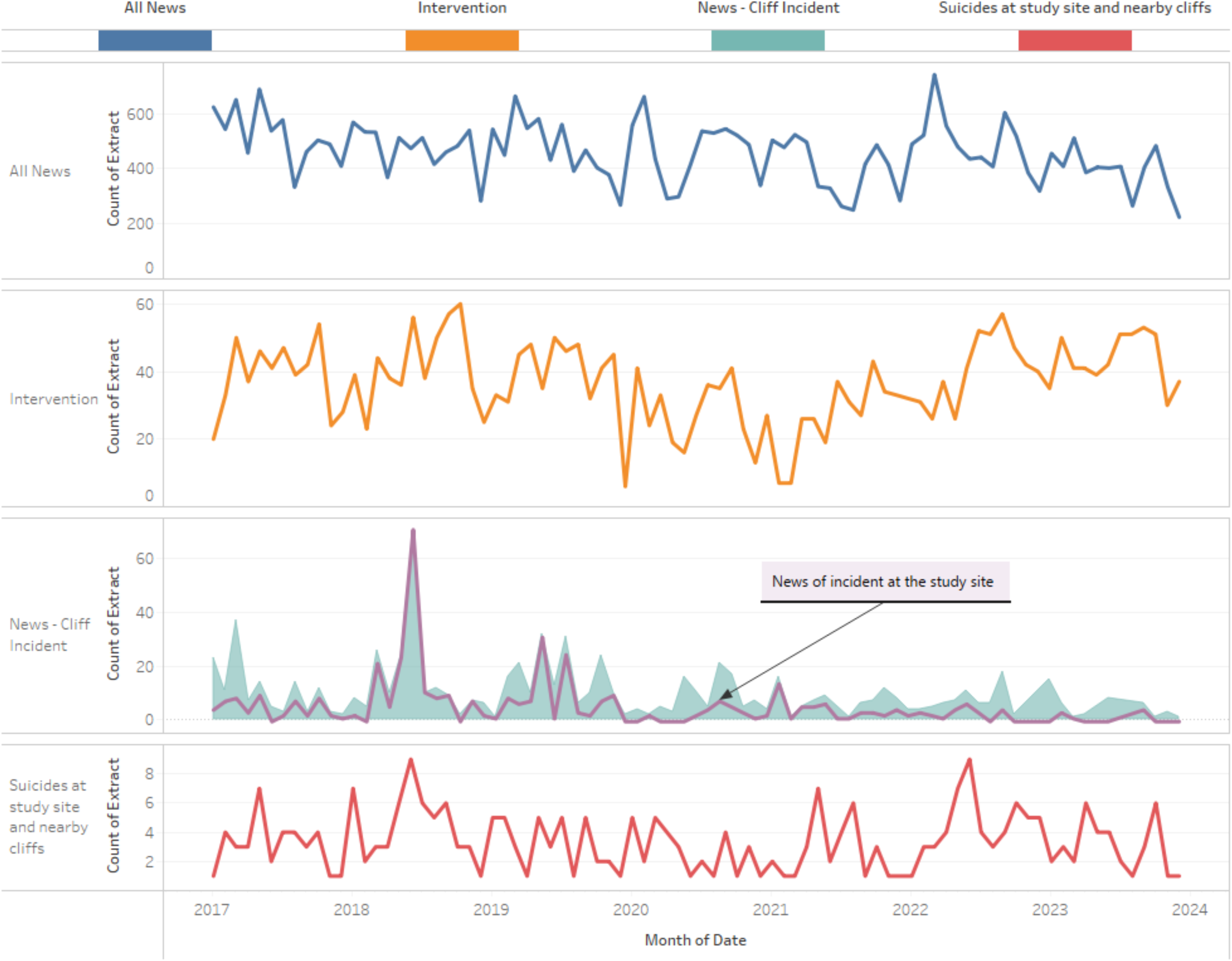
Monthly count of UK suicide news (by all methods), cliff suicide news (including specifically of suicides and attempts at the study site), interventions and suspected suicides at the study site and immediately adjacent cliffs (2017-2023)

We tested bidirectional lagged relationships between media reports of suicidal behaviour and suicide-related incidents over the study period (please see online Appendices for full details). Table 2 and 3 summarise, respectively, significant and non-significant associations between daily and weekly news reports and suspected suicides at the study site and immediately adjacent cliffs (n=278), and crisis interventions at the site (N=3,050). Please note that, even when reaching the threshold of statistical significance (p<0.05), all tested correlations were ‘weak’ or ‘very weak’ (r<0.4), except for the ‘moderate (0.4<r<0.59) association between daily news (by all methods) and interventions.

**Table 2.** Associations between news reports and suspected suicides at the study site and immediately adjacent cliffs (N=278) (2017-2023).

| Significant Associations ($p<0.05$) | r range |
| --- | --- |
| **<u>Daily coverage</u>** |  |
| Higher volume of **all UK suicide news (by all methods)**<br>→ Increase in suicides 1, 2, 7 and 9 days later | .05 |
| Higher volume of **cliff suicide news**<br>→ Increase in suicides on the same day and a day later | .09-.11 |
| Higher volume of **study site and nearby cliffs suicide news**<br>→ Increase in suicides on the same day, 1, 2 and 6 days later | .05-.13 |
| Higher volume of **study site suicide news (excluding nearby cliffs)**<br>→ Increase in suicides the same day, 1 and 2 days later | .06-.14 |
| <u>Weekly coverage</u> |  |
| More cliff suicides news<br>→ Increase in suicides on the same week | .24 |
| More study site and nearby cliffs suicide news<br>→ Increase in suicides on the same week and a week later | .11-.27 |
| More study site suicide news (excluding nearby cliffs)<br>→ Increase in suicides on the same week and a week later | .11-.27 |
| Non-significant associations |  |
| Weekly volume of all UK suicide news (by all methods) |  |

**\*Bolded headline types were found in Granger causality tests to significantly predict values of suicide in the future.**

### News Headlines with Explicit Method and/or Location Details

We repeated these analyses with a focus on news stories of suicides at all cliff locations and at the study site (and nearby cliffs) where explicit method and/or location details were included in the headline (as a proxy for poor quality reporting). The results of these tests are summarised in Table 4 below (see online Appendix for full details). Once again, statistical associations were found to be weak or very weak.

**Table 3.**
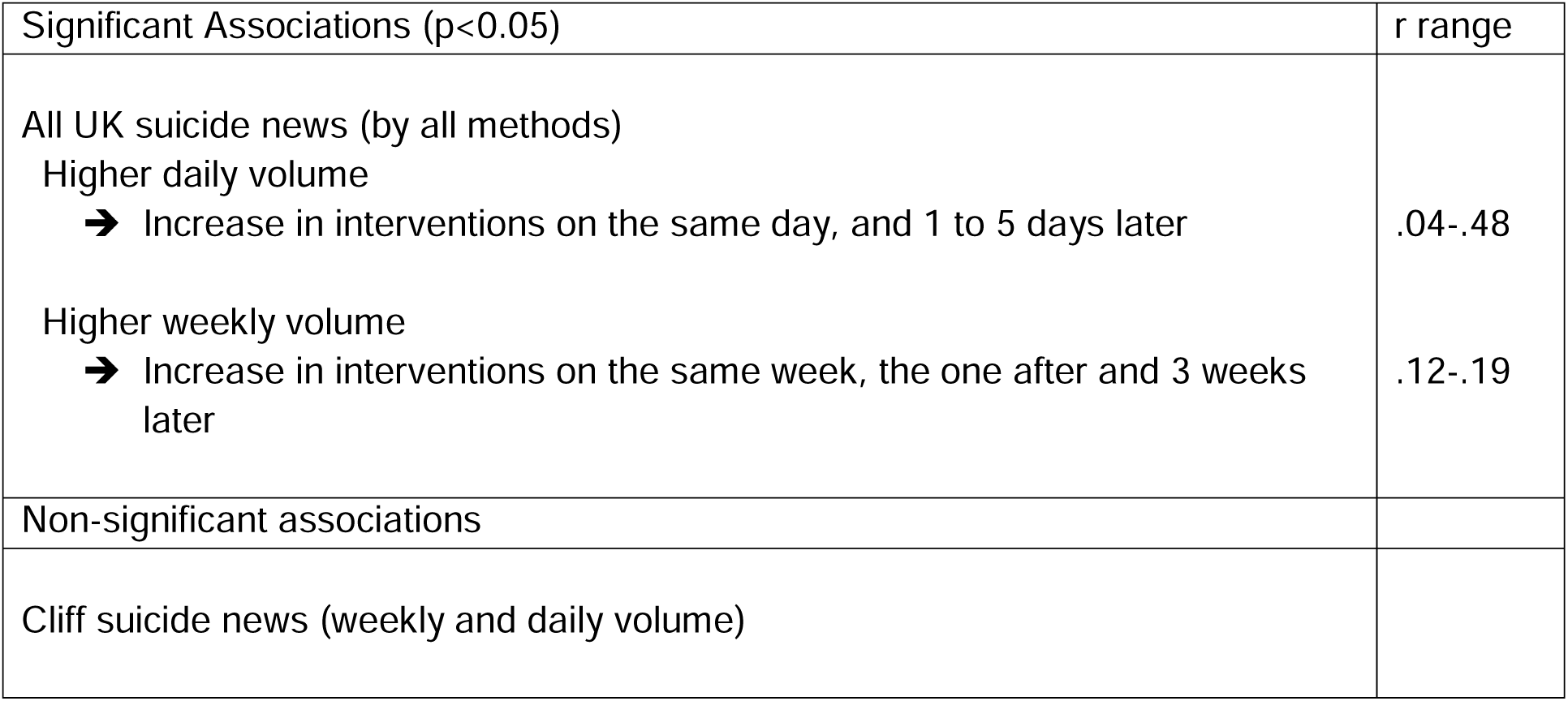
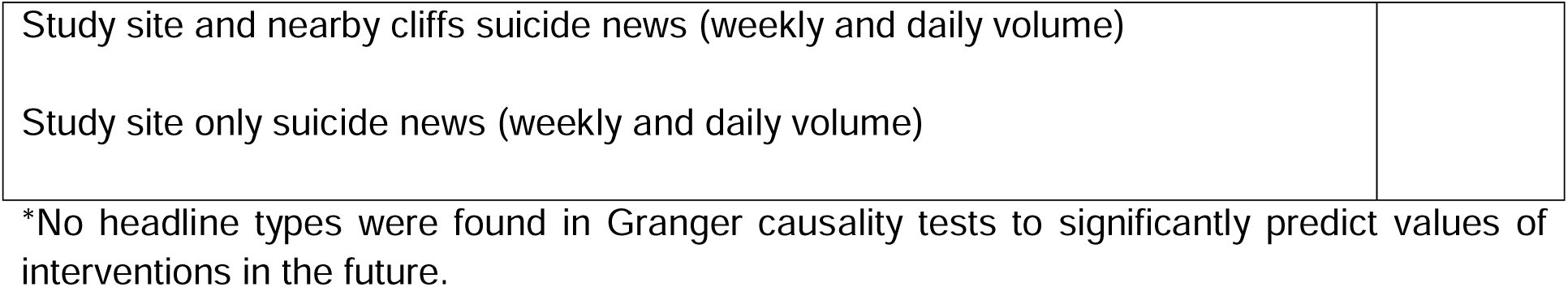
Associations between news reports and interventions at the study site (N=3,050) (2017-2023).

| Significant Associations (p<0.05) | r range |
| --- | --- |
| All UK suicide news (by all methods) |  |
| Higher daily volume<br>→ Increase in interventions on the same day, and 1 to 5 days later | .04-.48 |
| Higher weekly volume<br>→ Increase in interventions on the same week, the one after and 3 weeks later | .12-.19 |
| Non-significant associations |  |
| Cliff suicide news (weekly and daily volume) |  |
| Study site and nearby cliffs suicide news (weekly and daily volume) |  |
| Study site only suicide news (weekly and daily volume) |  |

\*No headline types were found in Granger causality tests to significantly predict values of interventions in the future.

**Table 4.** Associations between news headlines featuring explicit method and/or location details and suicide-related incidents (2017-2023).

| Significant Associations ($p < 0.05$) | r range |
| --- | --- |
| **Daily news all cliff suicide (n=588)**<br>→ Increase in suspected suicides on the day and a day later | .10-.12 |
| **Daily news of suicides at the study site and nearby cliffs (n=350)**<br>→ Increase in suspected suicides on the same day, 1, 2 and 6 days later | .05-.14 |
| Non-significant associations |  |
| Daily volume of headlines on suicides at all cliff locations, and specifically at the study site and nearby cliffs<br>→ No significant association with crisis interventions |  |

\***Bolded headline types were found in Granger causality tests to significantly predict values of suicide in the future.**

### Repeat Coverage of Suicides at the Study Site

Between 1^st^ January 2017 and 31^st^ December 2023, there were six weeks when stories of suicides at the study site featured in the news ten times or over (range: 10-48). These included two consecutive weeks in 2018 (total reports=58) and a further four weeks between March 2018 and July 2019 (with 16, 17, 26 and 18 weekly reports). In contrast, there were never more than four weekly reports in 2020, 2022 and 2023 or five in 2021. In 2017, a single week saw the publication of seven stories relating to suicidal behaviour at the study site, and another of eight news reports.

This pattern of repeat reporting over individual weeks reflects the repeat coverage of specific incidents (in two cases murder-suicides) at the study site, and related Coroner’s inquiries. In total, five incidents featured in the news over 10 times (range: 15 to 59 news items relating to a single incident, total news items=140, 34.7% of all study site news in this period). All of these 140 news stories were published in 2018 (n=85) and 2019 (n=55). In subsequent years, no individual was named in news of suicides at the study site more than three times (except for a woman named in seven reports in 2021).

After 2017 and 2018 (when respectively 2 and 24 news stories reported on multiple, apparently disconnected suicides at the study site (e.g. *“Ten deaths in a fortnight at beauty spot”*), there were also no headlines making explicit reference to multiple deaths at the site.

Given the marked reduction in repeat coverage of suicides at the study site from 2020 (both multiple reports of a single incident, and single news items reporting on multiple incidents), and the particularly high volume of reporting in 2018-19 (see Table 1), we conducted a separate analysis of the relationship between daily news reports of suicides at the study sites and incidents at the location in 2018-19, and then during 2020-2023. A summary of results is displayed in Table 5 below (please see online Appendix for further detail), whilst Figure 3 below displays the weekly volume of suicide news, suspected suicides and interventions relating specifically to the study site between 5^th^ March 2018 and 7^th^ July 2019,

**Figure 3.**
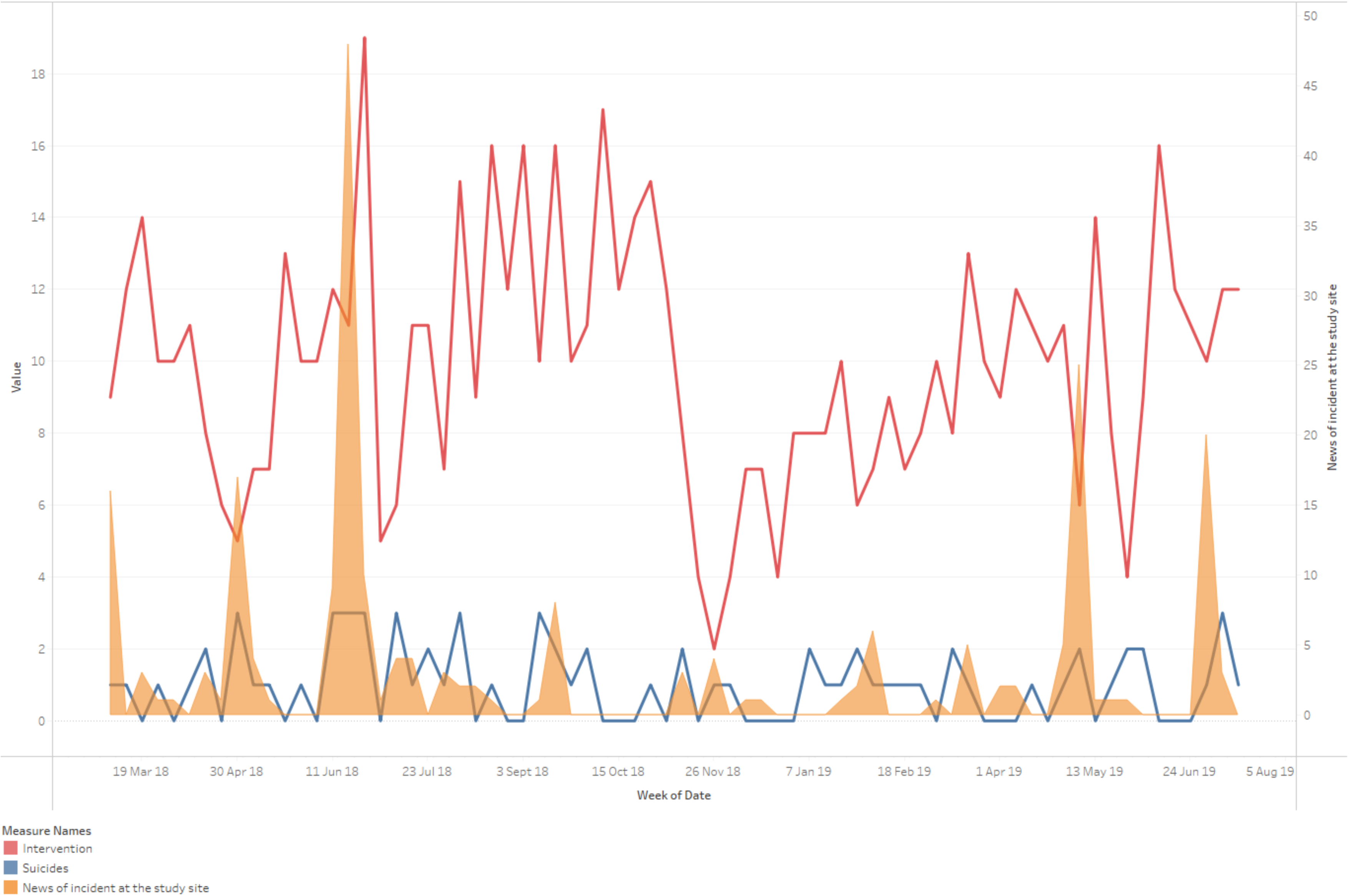
Weekly count of study site suicide news, suspected suicides at the location and immediately adjacent cliffs, and crisis interventions between 5^th^ March 2018 and 7^th^ July 2019.

**Table 5.** Associations between daily news reports of suicidal behaviour at the study sites and incidents at the location (2018-2019 vs. 2020-2023).

| 2018-2019 | r range |
| --- | --- |
| More daily reports of suicides at the study site (n=247)<br>➔ **Significant increase in suicides at the study site and immediately adjacent cliffs on the day and one, two and six days later\***<br>➔ No association with crisis interventions | .09-.18 |
| 2020-2023 |  |
| More daily reports of suicides at the study site (n=103)<br>➔ Significantly more suicides at the study site and immediately adjacent cliffs on the same day only<br>➔ No association with crisis interventions | .18 |

**\*Daily values of news stories of suicides at the study site in 2018-2019 were found in Granger causality tests to significantly predict values of suicide in the near future.**

the period with the highest volume of reporting (n=221, 54.8% of all study site news stories between 2017 and 2023) and repeat coverage.

## Discussion

As reflected in current WHO guidance for media professionals, several studies have shown that “*media reporting of suicide can lead to subsequent increases in suicidal behaviours … related to the amount and prominence of media coverage, with repeated coverage and high-impact stories being most strongly associated with imitative behaviours. […] Overt description of suicide by a particular method often leads to increases in suicidal behaviour employing that method*” [6]. This effect has been found to be especially marked for reports of celebrity suicides [2], with fewer studies exploring the impact of news coverage of suicides at well-known locations. We therefore investigated the relationship between news coverage of suicide and incidents at a high-frequency location in the UK, with an average of almost 40 suicides per year (over the past seven years).

Using data from the Samaritans’ media monitoring database between 1^st^ January 2017 and 31^st^ December 2023, we found that reports of suicides at the study site and other cliff locations represent a small proportion of all UK news pertaining to suicides and suicide attempts (by all methods). Overall, reports of suicidal behaviour at this specific location represented 1% of all UK news coverage of suicide between 2017 and 2023, and around 50% of all reports relating to cliff suicides. Of note is the decreasing volume of news stories focusing on cliff suicides and specifically deaths at the study site (e.g. there were eight news stories of suicides at this location in 2023, versus a peak of 151 in 2018), and the decrease in repeat coverage of specific incidents in 2020-2023 compared to earlier years (50% of all news stories pertaining to the study site in 2017-2019 (150/300) focused on six specific incidents, including two murder-suicides featuring, respectively, in 32 and 59 individual news items). However, the proportion of headlines including explicit reference to method and/or location details remained high throughout the study period (featuring, on average, in 74.5% of all cliff suicide news stories and 80.9% of all news relating to suicides at the study site).

Our analysis suggests statistically significant but very small correlations between suicides at the study site and immediately adjacent cliffs and reports of a) all suicide news, b) all cliff news, and c) site-specific news (except for weekly coverage of all suicide news), with the latter predicting daily values of subsequent suicides (generally within a day or two, and no more than 9 days later). These associations were slightly stronger (but nonetheless ‘weak’ to ‘very weak’ in statistical terms) on a weekly (as opposed to daily) basis; for reports focusing specifically on suicides at the study site and other cliffs (as opposed to all suicide methods); and in 2018-19, the years with the greatest volume of reporting and repeat coverage.

In contrast, news of suicides at the study site was not associated with the number of suicides in the immediate aftermath of reporting in 2020-2023, when there were fewer reports, fewer stories being covered in the news on multiple occasions, and no headlines making explicit reference to multiple deaths at the site.

Contrary to our predictions, sub-analyses focusing on news stories that included method and/or location details in the headline did not have a significantly stronger association with incidents at the study site. Their relationship, albeit statistically significant, was also ‘very weak’.

Associations between news reports and crisis interventions at the study site followed a somewhat different pattern:

1. This relationship was only significant for daily and weekly news coverage of all suicides (irrespective of the reports’ suicide methods or locations), and strongest on days of increased reporting (reaching a ‘moderate’ level) – but the volume of suicide news did not significantly predict values of future interventions at the location.
2. Reporting of cliff suicides and incidents at the study site was not significantly associated with the number of interventions at the site – even when only taking into account data from 2018 and 2019 (the years with the greatest volume of reporting and repeat coverage).

### Strengths and Limitations of the Research

Our findings are based on a well-established, evidence-informed media monitoring database, which captures media reports of suicides and attempted suicides in the UK [12]. This was carefully adapted for use in this study. However, our database may not capture all relevant reports or all important aspects of reporting. Also, our focus did not extend to broadcasts or other media formats, and we did not estimate the impact or reach of different news reports or *types* of reports (including the circulation rate of different story types, in news as well as social media channels). The nature and ‘newsworthiness’ of different incidents being reported at different times (e.g. a ‘murder-suicide’ or the death of a well-known individual) are likely confounders for the volume and quality of associated news coverage.

The conclusions that may be drawn based on this analysis may also be limited by the speed at which trends in media (especially ‘new’ media) change, and the complex and dynamic ways in which their influence and effects manifest themselves in relation to different audiences and individuals. These were beyond the scope of the current investigation but would benefit from further research.

In addition, our analyses were based on *suspected* suicides. Although research suggests that suspected suicide data from established real-time surveillance systems can accurately reflect suicide rates confirmed by official statistics [13], any inferences or conclusions about *suicides* should be made cautiously. Similarly, not all individuals presenting in distress at the study site may be captured in the crisis intervention database we analysed, so the possibility of missing data should also be taken into account (but with no reason to believe this would follow a non-random pattern and therefore introduce significant bias in the analysis).

### Implications and Recommendations

The factors influencing increases – or indeed decreases – in suicides and interventions at specific locations (and more generally) are known to be complex, dynamic and multi-faceted. Therefore, it is perhaps unsurprising that the observed media effects were very small – particularly over what was a fairly long timeframe, affected by significant events such as the Covid-19 pandemic and associated restrictions, and subsequent cost of living crisis. A recent meta-analysis of 31 studies examining the association between reporting on suicides and subsequent suicides in the general population found that news of celebrity deaths had “a meaningful impact on total suicides in the general population”, which was “larger for increases by the same method as used by the celebrity”. However, “general reporting of suicide did not appear to be associated with suicide, although associations for certain types of reporting cannot be excluded” [2].

Our finding that media reports of suicide-related incidents can predict (however modestly) increases in suicide underscores the importance of continued efforts to monitor and improve the quality of news and other media reports of suicide. This remains a key priority in national efforts to reduce suicides [14] and may include reducing sensationalist, detailed and repeated coverage of suicides, and ensuring that articles relating to suicides include messages of hope and recovery, and signposting to help and support [15].

Our data suggests some marked improvements in the reporting of deaths at this high-frequency location in recent years (specifically in the volume of such reports, and reduction in repeat coverage), but also some areas which would benefit from further attention (notably the continued use of exact and explicit location details in headlines).

Further research is needed for a more nuanced understanding of the impact of different news reports – and wider stories/narratives – surrounding suicides and the study site in different spaces, including in online and ‘pro-choice’ spaces [16]. For example, research could cast light on whether specific narratives that may deter individuals from specific methods or locations (such as concern about the impact of suicide on others and fear of survival with injuries [17]) in news stories could dissuade people from choosing the location as a site/method of suicide. How ‘mainstream’ news stories circulate through various social media channels, their reach and impact, is another potential area of research.

Qualitative research with people with lived/living experience could provide useful means to explore these questions. For example, the perspectives of people who have travelled or considered travelling from afar to attempt suicide at the study site could offer some crucial insights into the ‘stories’ attracting them to this location, the origin of these stories, and the role of the news and other media in ‘implanting’, reinforcing and/or challenging them. In turn, this could inform a more nuanced analysis of the impact of different types of news stories/messages on incidents at this high-frequency site, and influence changes in reporting and communications to reduce the cognitive availability and ‘attractiveness’ of coastal methods/locations of suicide.

## Conclusions

This study is the first systematic analysis of the relationship between news coverage of suicide and suicidal behaviour at a high-frequency location. Our results, based on data collected over seven years, suggest that there is a small but significant association between daily and weekly news coverage of all suicides (irrespective of the reports’ suicide methods or locations) and crisis interventions at the study site (particularly on the same day), and between volume of reporting and increases in suspected suicides in the immediate aftermath of reporting (generally within a day or two, and no more than nine days later). The latter association was more marked (albeit statistically ‘weak’) in 2018-19, the years with the greatest volume of reporting and repeat coverage of ‘high-impact’ stories (e.g. reports of ‘murder-suicides’), and following coverage of method– and location-specific suicides (i.e. of incidents at cliff locations and at the study site, vs. reporting of suicides by all methods). In contrast in 2020-23, when there were fewer reports of suicides at the study site, fewer such stories being covered in the news on multiple occasions, and no headlines making explicit reference to multiple deaths at the site, this association was no longer significant. These findings underscore the importance of responsible portrayal of suicide in the news and other media.

## Declarations

### Ethical Approval and Consent to Participate

The study was scrutinised and approved by the Psychology Department Research Ethics Committee at Middlesex University (ref: 8412), subject to the terms of a letter of agreement with the Public Health Directorate of the participating Suicide Prevention Partnership and Memorandum of Understanding with Samaritans.

## Consent for Publication

Not applicable.

## Availability of Data and Materials

Currently data access is restricted, although aggregate anonymised data collected could be made available with the permission of the participating Suicide Prevention Partnership.

## Authors’ Contributions

LM: conceptualization, funding and data acquisition, methodology, project administration, writing. RS: data curation, methodology, formal analysis, writing. IM: conceptualization, funding and data acquisition. AB: data coding and curation. IK: data visualisation and analysis.

All authors reviewed and edited the final manuscript.

## Acknowledgments

The research team would like to thank the Public Health Directorate and Operational Response Unit of the participating County Council for all their support throughout the research. We are also very grateful to Lorna Fraser, Monica Hawley and Rachel Davidson from the Samaritans’ Media Advisory Team.

