## Supplementary material for "How does news coverage of suicide affect suicidal behaviour at a high-frequency location? A seven-year time-series analysis": Online appendix

### A. Associations between News Reports and Suspected Suicides at the Study Site and Immediately Adjacent Cliffs (N=278), between 1st January 2017 and 31st December 2023

**A1. NEWS COVERAGE OF ALL SUICIDES (BY ALL METHODS)**

***Are daily media reports of all suicides (N=38,595) related to suspected suicides?***

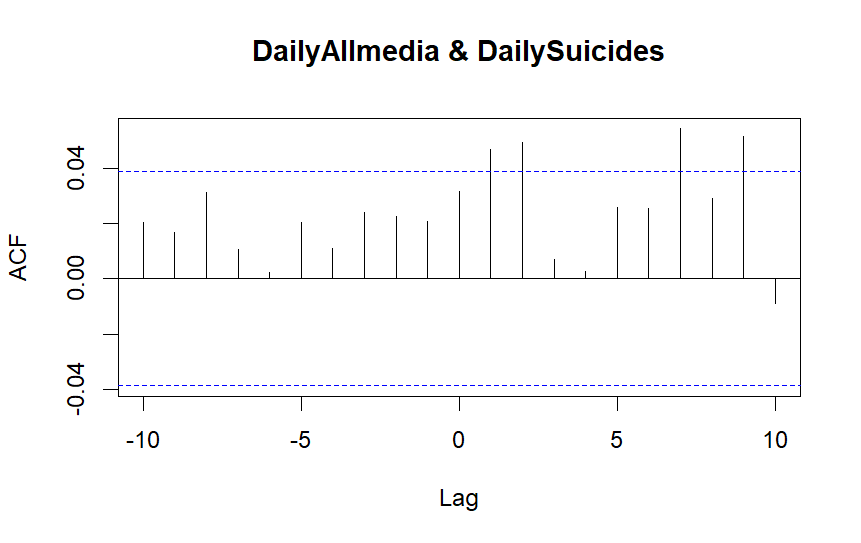

An increase in headlines is associated with an increase in suicides one day (r = .05), two days (r = .05), seven days (r = .05) and nine days (r =.05) later.

Correlations by lag

| **Lag** | 0 | 1 | 2 | 3 | 4 | 5 | 6 | 7 | 8 | 9 | 10 |
| --- | --- | --- | --- | --- | --- | --- | --- | --- | --- | --- | --- |
| **Correlation** | .03 | .05 | .05 | .01 | .00 | .03 | .03 | .05 | .03 | .05 | -.01 |

Values of headlines Granger cause values of suicides in the future (F = 4.22, *p* = .04); suicides do not Granger cause headlines (F = 1.11, *p* = .29).

***Are weekly media reports of all suicides (N=38,595) related to suspected suicides?***

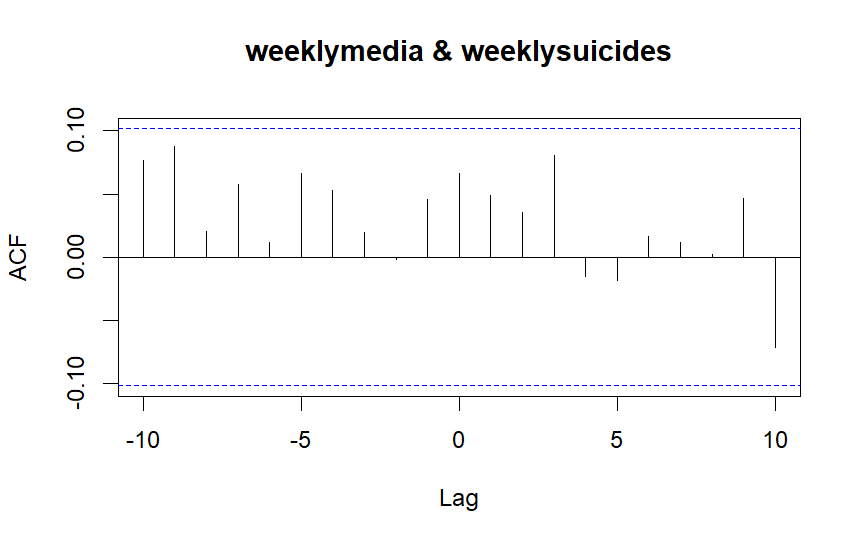

An increase in headlines is not associated with an increase in suicides.

Correlations by lag

| **Lag** | -5 | -4 | -3 | -2 | -1 | 0 | 1 | 2 | 3 | 4 | 5 |
| --- | --- | --- | --- | --- | --- | --- | --- | --- | --- | --- | --- |
| **Correlation** | .07 | .05 | .02 | .00 | .05 | .07 | .05 | .04 | .08 | -.02 | -.02 |

Values of headlines do not Granger cause values of suicides in the future (F = 0.18, *p* = .67); suicides do not Granger cause headlines (F = 0.74, *p* = .39).

**A2. NEWS COVERAGE OF ALL CLIFF SUICIDES**

***Are daily media reports of all cliff suicides (N=789) related to suspected suicides?***

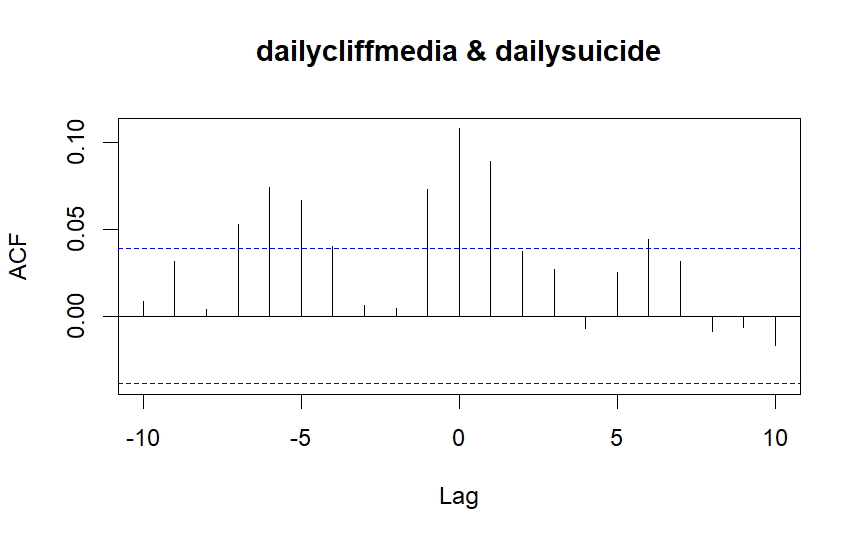

An increase in headlines is associated with an increase in suicides on the day (r = .11), one day later (r = .09) and six days later. An increase in suicides is associated with an increase in headlines a day later (r = .07) and five to seven days later.

Correlations by lag

| **Lag** | -7 | -6 | -5 | -4 | -3 | -2 | -1 | 0 | 1 | 2 | 3 | 4 | 5 |
| --- | --- | --- | --- | --- | --- | --- | --- | --- | --- | --- | --- | --- | --- |
| **Correlation** | .05 | .07 | .07 | .04 | .01 | .00 | .07 | .11 | .09 | .04 | .03 | -.01 | .03 |

Values of headlines Granger cause values of suicides in the future (F = 6.35, *p* = .01); suicides Granger cause future values of headlines (F = 13.93, *p* < .001).

***Are weekly media reports of all cliff suicides (N=789) related to suspected suicides?***

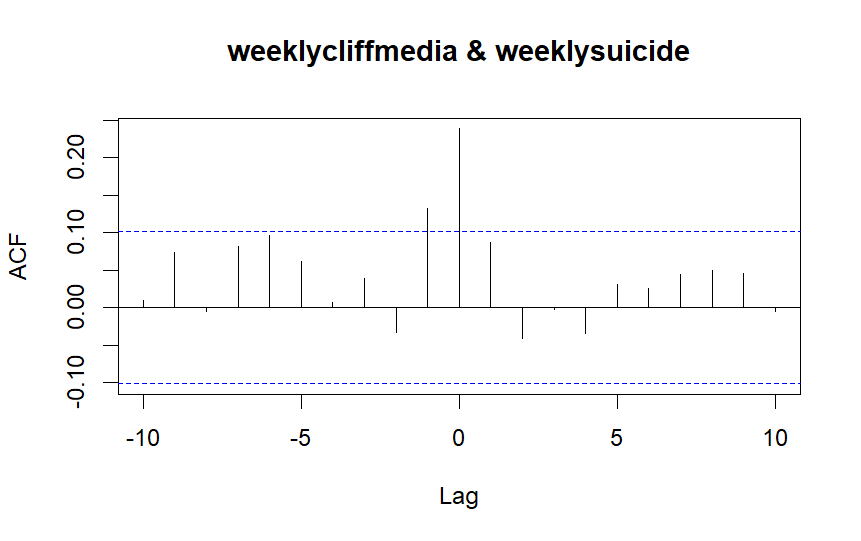

An increase in headlines is associated with an increase in suicides the same week (r = .24). An increase in suicides is associated with an increase in headlines a week later (r = .13).

Correlations by lag

| **Lag** | -10 | -9 | -8 | -7 | -6 | -5 | -4 | -3 | -2 | -1 | 0 | 1 |
| --- | --- | --- | --- | --- | --- | --- | --- | --- | --- | --- | --- | --- |
| **Correlation** | .01 | .07 | -.01 | .08 | .10 | .06 | .01 | .04 | -.03 | .13 | .24 | .09 |

Values of headlines do not Granger cause values of suicides in the future (F = 0.64, *p* = .43); suicides Granger cause future values of headlines (F = 6.48, *p* = .01).

**A3. NEWS COVERAGE OF SUICIDES AT THE STUDY SITE AND IMMEDIATELY ADJACENT CLIFFS**

***Are daily media reports of suicides at the study site and immediately adjacent cliffs (N=433) related to suspected suicides?***

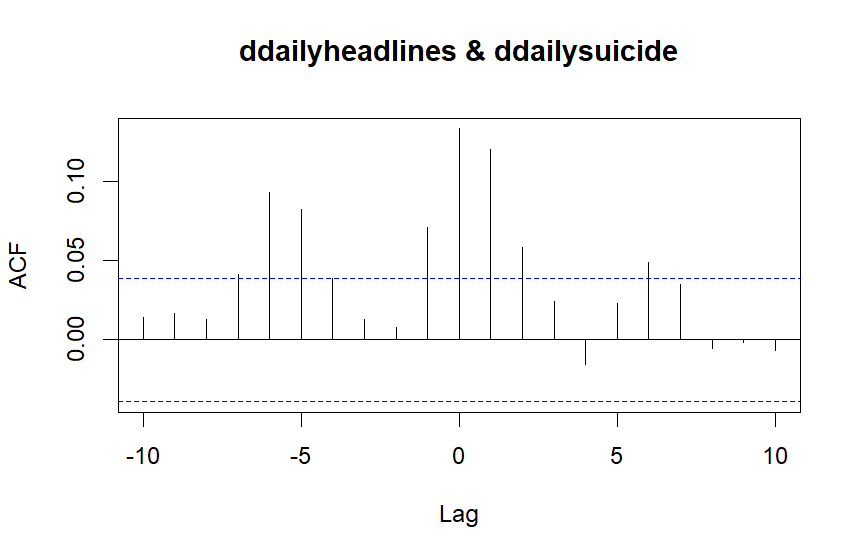

An increase in headlines is associated with an increase in suicides on the day (r = .13), one day (r = .12), two days (r = .06) and six days (r = .05) later. An increase in suicides is associated with an increase in headlines a day later (r = .07) and five to six days later.

Correlations by lag

| **Lag** | -6 | -5 | -4 | -3 | -2 | -1 | 0 | 1 | 2 | 3 | 4 | 5 | 6 |
| --- | --- | --- | --- | --- | --- | --- | --- | --- | --- | --- | --- | --- | --- |
| **Correlation** | .09 | .08 | .04 | .01 | .01 | .07 | .13 | .12 | .06 | .02 | -.02 | .02 | .05 |

Daily values of headlines Granger cause values of suicides in the future (F = 10.84, *p* < .001); suicides Granger cause future daily values of headlines (F = 13.15, *p* < .001).

***Are weekly media reports of suicides at the study site and immediately adjacent cliffs (N=433) related to suspected suicides?***

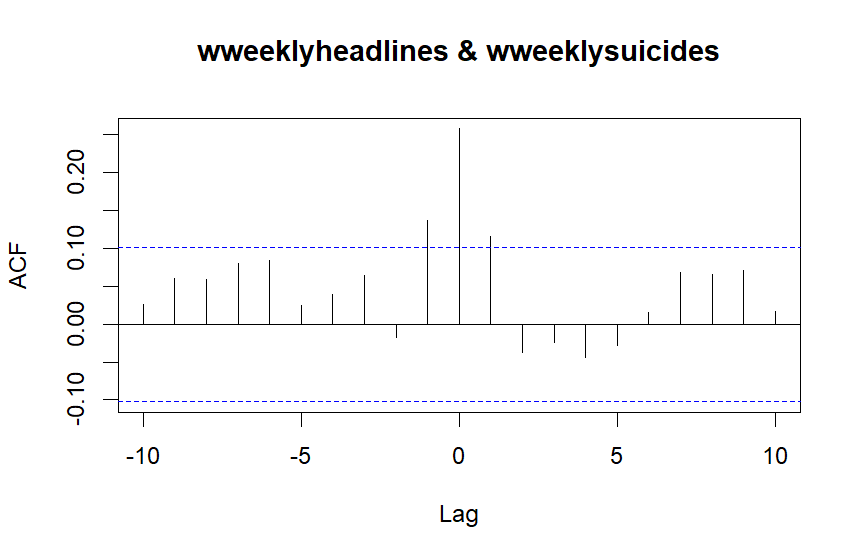

An increase in headlines is associated with an increase in suicides the same week (r = .27), and a week later (r = .11). An increase in suicides is associated with an increase in headlines a week later (r = .14).

Correlations by lag

| **Lag** | -7 | -6 | -5 | -4 | -3 | -2 | -1 | 0 | 1 | 2 | 3 | 4 | 5 |
| --- | --- | --- | --- | --- | --- | --- | --- | --- | --- | --- | --- | --- | --- |
| **Correlation** | .08 | .08 | .02 | .04 | .07 | -.02 | .14 | .26 | .12 | -.04 | -.02 | -.05 | -.03 |

Weekly values of headlines do not Granger cause values of suicides in the future (F = 1.50, *p* = .22); weekly suicides Granger cause future weekly values of headlines (F = 7.02, *p* = .008).

**A4. NEWS COVERAGE OF SUICIDES AT THE STUDY SITE (EXCLUDING IMMEDIATELY ADJACENT CLIFFS)**

***Are daily media reports of suicides at the study site (N=403) related to suspected suicides?***

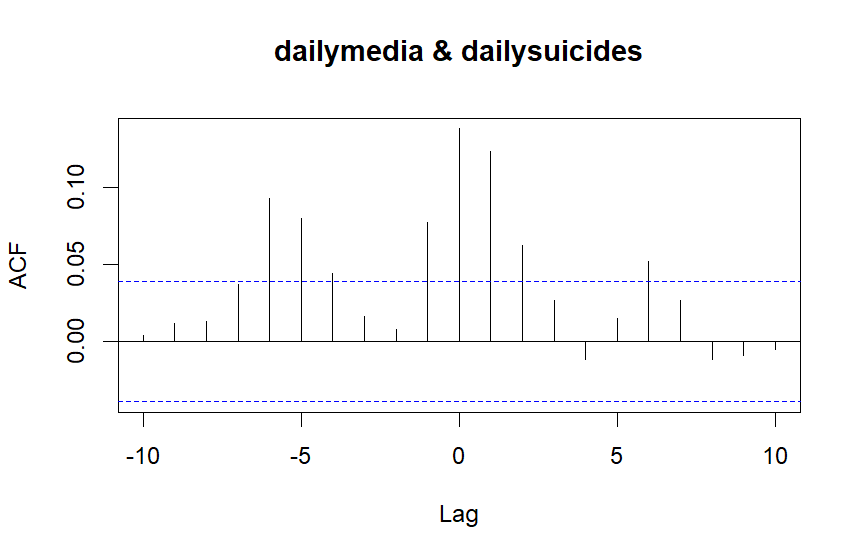

An increase in headlines is associated with an increase in suicides on the day (r = .14), one day (r = .12), two days (r = .06) and six days later. An increase in suicides is associated with an increase in headlines a day later (r = .08) and five to six days later.

Correlations by lag

| **Lag** | -7 | -6 | -5 | -4 | -3 | -2 | -1 | 0 | 1 | 2 | 3 | 4 | 5 |
| --- | --- | --- | --- | --- | --- | --- | --- | --- | --- | --- | --- | --- | --- |
| **Correlation** | .04 | .09 | .08 | .04 | .02 | .01 | .08 | .14 | .12 | .06 | .03 | -.01 | .02 |

Daily values of headlines Granger cause values of suicides in the future (F = 10.98, *p* < .001); suicides Granger cause future daily values of headlines (F = 15.60, *p* < .001).

***Are weekly media reports of suicides at the study site (N=403) related to suspected suicides?***

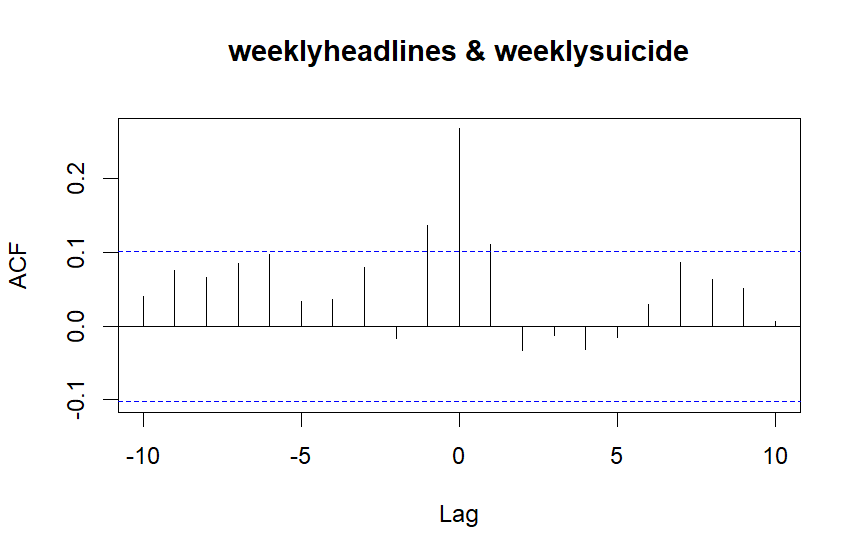

An increase in headlines is associated with an increase in suicides the same week (r = .27), and a week later (r = .11). An increase in suicides is associated with an increase in headlines a week later (r = .14).

Correlations by lag

| **Lag** | -7 | -6 | -5 | -4 | -3 | -2 | -1 | 0 | 1 | 2 | 3 | 4 | 5 |
| --- | --- | --- | --- | --- | --- | --- | --- | --- | --- | --- | --- | --- | --- |
| **Correlation** | .09 | .10 | .03 | .04 | .08 | .02 | .14 | .27 | .11 | -.03 | -.01 | -.03 | -.02 |

Weekly values of headlines do not Granger cause values of suicides in the future (F = 1.07, *p* = .30); weekly suicides Granger cause future weekly values of headlines (F = 7.04, *p* = .008).

### B. Associations between News Reports and Crisis Interventions at the Study Site (N=3,050), between 1st January 2017 and 31st December 2023

**B1. NEWS COVERAGE OF ALL SUICIDES (BY ALL METHODS)**

***Are daily media reports of all suicides (N=38,595) related to interventions?***

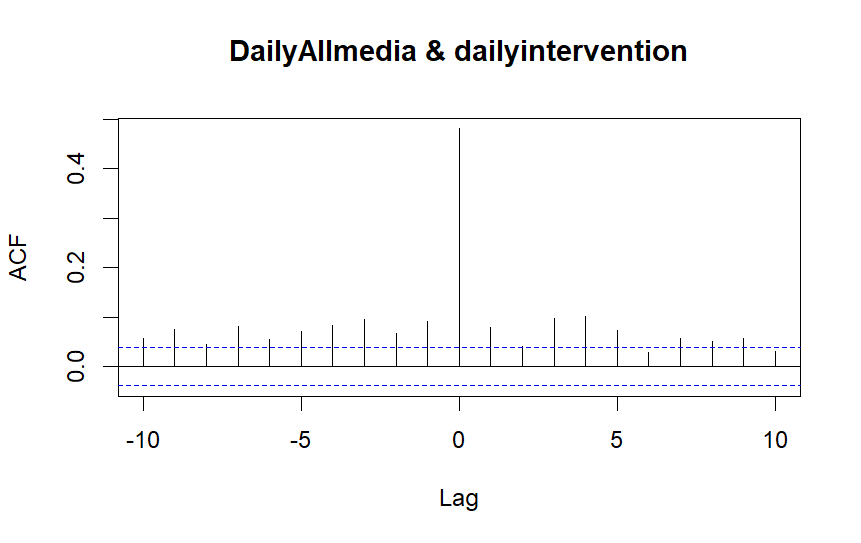

An increase in headlines is associated with an increase in interventions on the day (r = .48), and one to nine days later. An increase in interventions is associated with an increase in headlines one to ten days later (see table).

Correlations by lag

| **Lag** | -5 | -4 | -3 | -2 | -1 | 0 | 1 | 2 | 3 | 4 | 5 |
| --- | --- | --- | --- | --- | --- | --- | --- | --- | --- | --- | --- |
| **Correlation** | .07 | .08 | .10 | .07 | .09 | .48 | .08 | .04 | .10 | .10 | .07 |

Values of suicide reports just miss significance in Granger causing values of interventions in the future (F = 3.33, *p* = .068); interventions do not Granger cause media stories (F = 2.02, *p* = .16).

***Are weekly media reports of all suicides (N=38,595) related to interventions?***

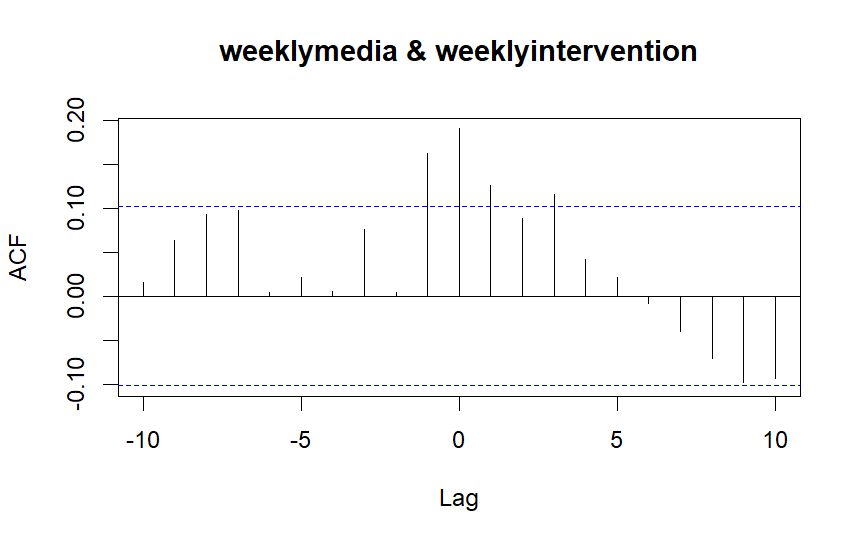

An increase in headlines is associated with an increase in interventions the same week (r = .19), one week (r = .13), and three weeks (r = .12) later. An increase in interventions is associated with an increase in headlines a week later (r = .16).

Correlations by lag

| **Lag** | -5 | -4 | -3 | -2 | -1 | 0 | 1 | 2 | 3 | 4 | 5 |
| --- | --- | --- | --- | --- | --- | --- | --- | --- | --- | --- | --- |
| **Correlation** | .02 | .01 | .08 | .01 | .16 | .19 | .13 | .09 | .12 | .04 | .02 |

Values of headlines do not Granger cause values of interventions in the future (F = 0.76, *p* = .38); the number of interventions do predict values of headlines in the future (F = 3.99, *p* = .046).

**B2. NEWS COVERAGE OF ALL CLIFF SUICIDES**

***Are daily media reports of all cliff suicides (N=789) related to interventions?***

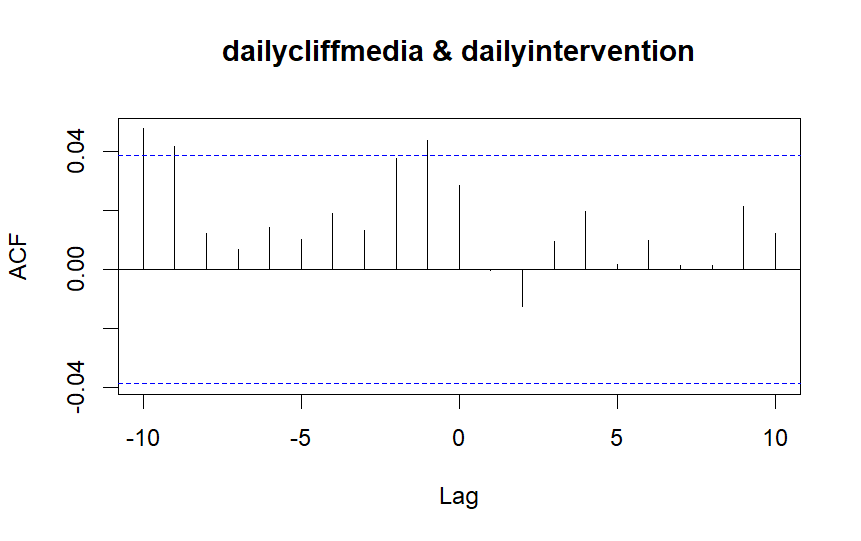

An increase in headlines is not associated with an increase in interventions. An increase in interventions is associated with an increase in headlines a day later (r = .07) and 9-10 days later.

Correlations by lag

| **Lag** | -10 | -9 | -8 | -7 | -6 | -5 | -4 | -3 | -2 | -1 | 0 | 1 | 2 |
| --- | --- | --- | --- | --- | --- | --- | --- | --- | --- | --- | --- | --- | --- |
| **Correlation** | .05 | .04 | .01 | .01 | .01 | .01 | .02 | .01 | .04 | .04 | .03 | .00 | -.01 |

Values of headlines do not Granger cause values of interventions in the future (F = 0.42, *p* = .52); interventions Granger cause future values of headlines (F = 4.16, *p* = .04).

***Are weekly media reports of all cliff suicides (N=789) related to interventions?***

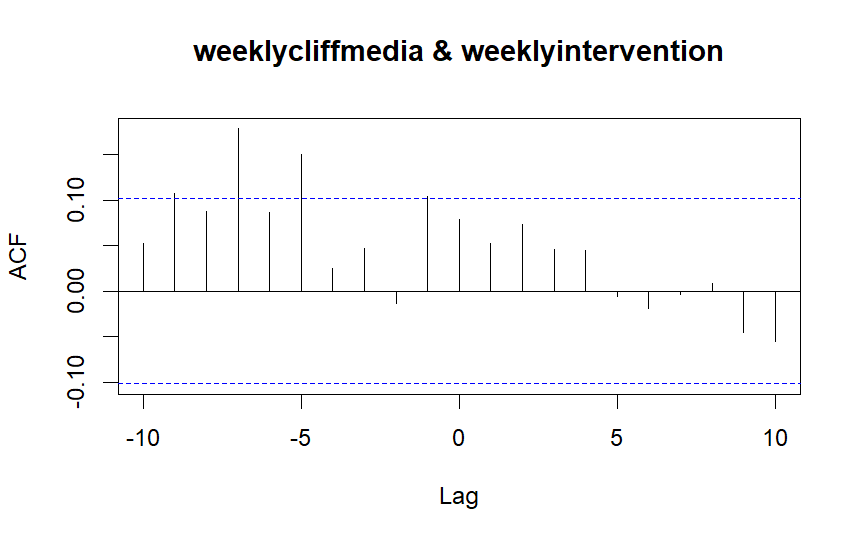

An increase in headlines is not associated with an increase in interventions. An increase in interventions is associated with an increase in headlines five and seven weeks later.

Correlations by lag

| **Lag** | -10 | -9 | -8 | -7 | -6 | -5 | -4 | -3 | -2 | -1 | 0 | 1 | 2 |
| --- | --- | --- | --- | --- | --- | --- | --- | --- | --- | --- | --- | --- | --- |
| **Correlation** | .05 | .11 | .09 | .18 | .09 | .15 | .03 | .05 | -.01 | .10 | .08 | .05 | .07 |

Values of headlines do not Granger cause values of interventions in the future (F = 0.55, *p* = .46); interventions do not Granger cause future values of headlines (F = 2.45, *p* = .12).

**B3. NEWS COVERAGE OF SUICIDES AT THE STUDY SITE AND IMMEDIATELY ADJACENT CLIFFS**

***Are daily media reports of suicides at the study site and immediately adjacent cliffs (N=433) related to interventions?***

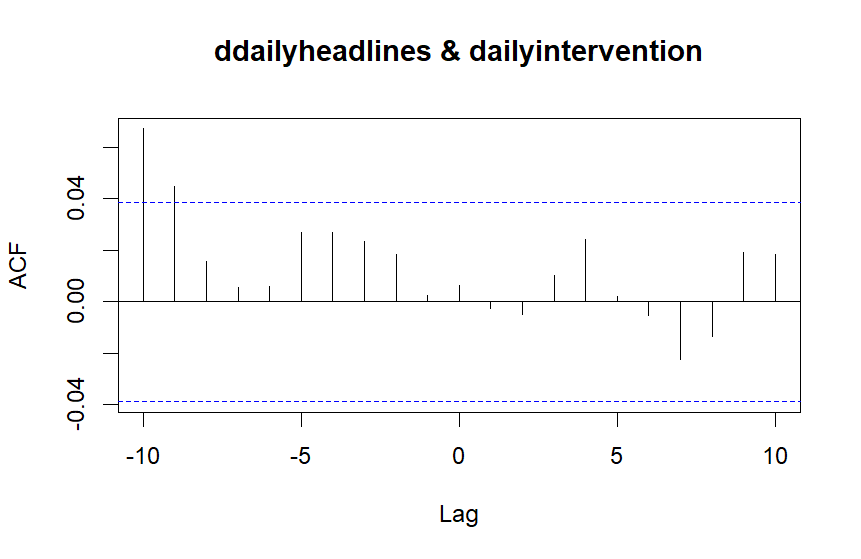

An increase in headlines is not associated with an increase in interventions. An increase in interventions is associated with an increase in headlines nine to 10 days later.

Correlations by lag

| **Lag** | -10 | -9 | -8 | -7 | -6 | -5 | -4 | -3 | -2 | -1 | 0 | 1 | 2 |
| --- | --- | --- | --- | --- | --- | --- | --- | --- | --- | --- | --- | --- | --- |
| **Correlation** | .07 | .05 | .02 | .01 | .01 | .03 | .03 | .02 | .02 | .00 | .01 | .00 | -.01 |

Values of headlines Granger do not cause values of interventions in the future (F = .10, *p* = .75); interventions do not Granger cause future values of headlines (F = .01, *p* = .94).

***Are weekly media reports of suicides at the study site and immediately adjacent cliffs (N=433) related to interventions?***

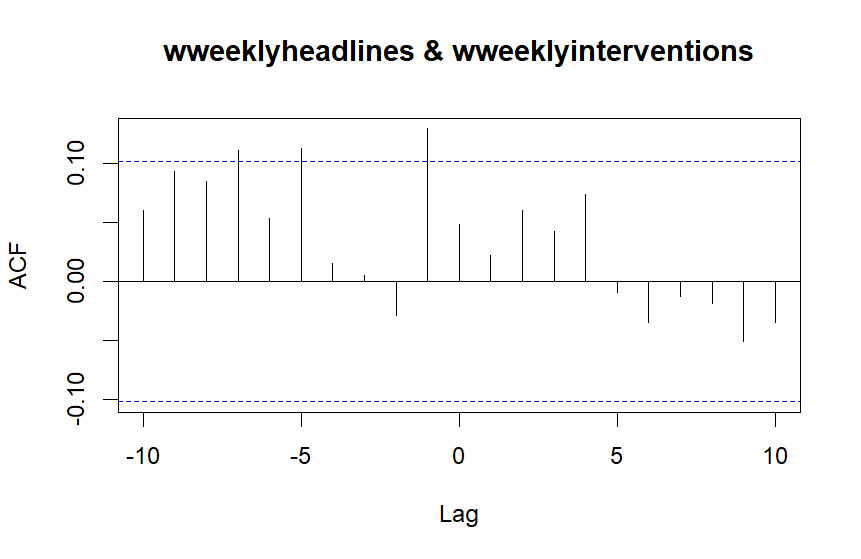

An increase in headlines is not associated with an increase in interventions. An increase in interventions is associated with an increase in headlines one week (r = .13), five weeks (r = .11) and seven weeks (r = .11) later.

Correlations by lag

| **Lag** | -10 | -9 | -8 | -7 | -6 | -5 | -4 | -3 | -2 | -1 | 0 | 1 | 2 |
| --- | --- | --- | --- | --- | --- | --- | --- | --- | --- | --- | --- | --- | --- |
| **Correlation** | .06 | .09 | .08 | .11 | .05 | .11 | .02 | .01 | -.03 | .13 | .05 | .02 | .06 |

Values of headlines do not Granger cause values of interventions in the future (F = .05, *p* = .82); interventions Granger cause future values of headlines (F = 5.44, *p* = .02).

**B4. NEWS COVERAGE OF SUICIDES AT THE STUDY SITE (EXCLUDING IMMEDIATELY ADJACENT CLIFFS)**

***Are daily media reports of suicides at the study site (N=403) related to interventions?***

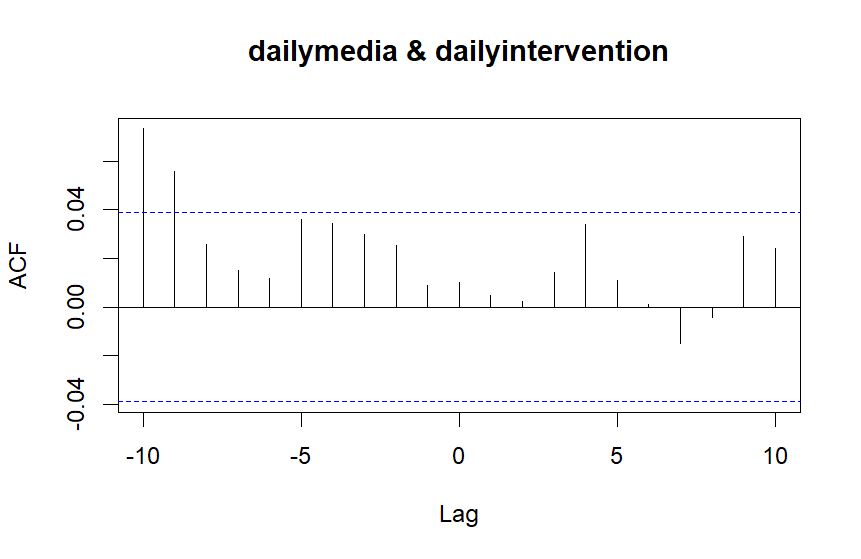

An increase in headlines is not associated with an increase in interventions. An increase in interventions is associated with an increase in headlines nine to 10 days later.

Correlations by lag

| **Lag** | -10 | -9 | -8 | -7 | -6 | -5 | -4 | -3 | -2 | -1 | 0 | 1 | 2 |
| --- | --- | --- | --- | --- | --- | --- | --- | --- | --- | --- | --- | --- | --- |
| **Correlation** | .07 | .06 | .03 | .02 | .01 | .04 | .04 | .03 | .03 | .01 | .01 | .01 | .02 |

Values of headlines do not Granger cause values of interventions in the future (F = .0002, *p* = .99); interventions do not Granger cause future values of headlines (F = .15, *p* = .70).

***Are weekly media reports of suicides at the study site (N=403) related to interventions?***

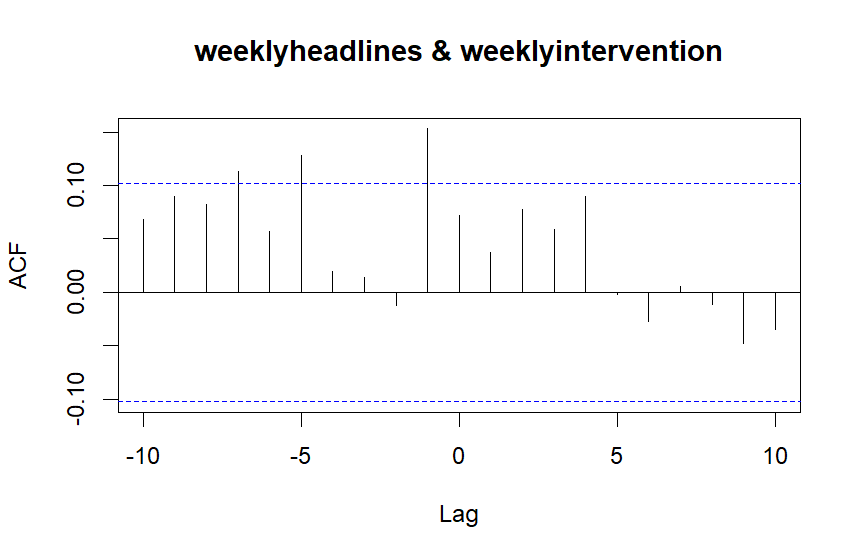

An increase in headlines is not associated with an increase in interventions. An increase in interventions is associated with an increase in headlines a week (r = .15), five weeks (r = .13) and seven weeks (r = .11) later.

Correlations by lag

| **Lag** | -10 | -9 | -8 | -7 | -6 | -5 | -4 | -3 | -2 | -1 | 0 | 1 | 2 |
| --- | --- | --- | --- | --- | --- | --- | --- | --- | --- | --- | --- | --- | --- |
| **Correlation** | .07 | .09 | .08 | .11 | .06 | .13 | .02 | .01 | -.01 | .15 | .07 | .04 | .08 |

Values of headlines do not Granger cause values of interventions in the future (F = .19, *p* = .66); interventions Granger cause future values of headlines (F = 7.01, *p* = .008).

### C. Associations between News Reports which Include Explicit Method and/or Location Details in the Headlines and Interventions and Suspected Suicides at the Study Site and Immediately Adjacent Cliffs (N=278), between 1^st^ January 2017 and 31^st^ December 2023

**C1. ALL CLIFF NEWS WITH METHOD AND/OR LOCATION DETAILS IN THE HEADLINE (N=588)**

***Are media stories of all cliff suicides where method and/or location are in the headline associated with suspected suicides at the study site and immediately adjacent cliffs?***

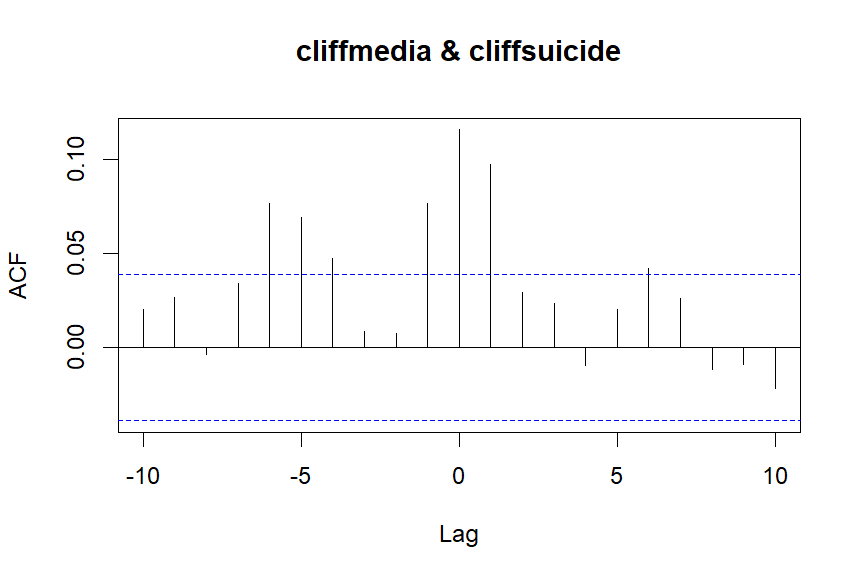

An increase in headlines was associated with an increase in suicides on the day (r = .12) and a day later (r = .10). An increase in suicides was associated with an increase in headlines a day later (r = .08) and four to six days later.

Correlations by lag

| **Lag** | -10 | -9 | -8 | -7 | -6 | -5 | -4 | -3 | -2 | -1 | 0 | 1 | 2 |
| --- | --- | --- | --- | --- | --- | --- | --- | --- | --- | --- | --- | --- | --- |
| **Correlation** | .02 | .03 | .00 | .03 | .08 | .07 | .05 | .01 | .01 | .08 | .12 | .10 | .03 |

Daily values of headlines Granger cause values of suicides in the future (F = 8.53, *p* = .004); suicides Granger cause future daily values of headlines (F = 15.44, *p* < .001).

***Are media stories of all cliff suicides where method and/or location are in the headline associated with interventions at the study site?***

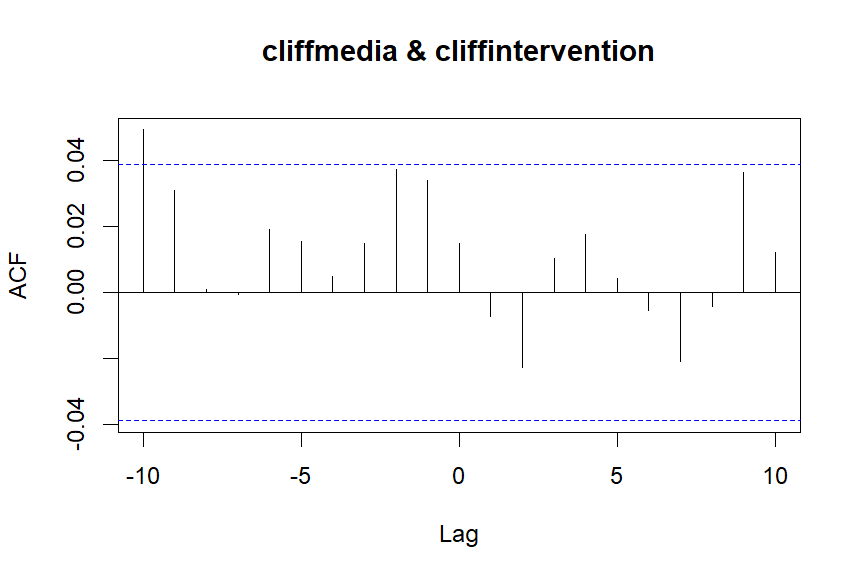

An increase in headlines was not associated with interventions. An increase in interventions was associated with an increase in headlines 10 days later (r = .05).

Correlations by lag

| **Lag** | -10 | -9 | -8 | -7 | -6 | -5 | -4 | -3 | -2 | -1 | 0 | 1 | 2 |
| --- | --- | --- | --- | --- | --- | --- | --- | --- | --- | --- | --- | --- | --- |
| **Correlation** | .02 | .03 | .00 | .03 | .08 | .07 | .05 | .01 | .01 | .08 | .12 | .10 | .03 |

Daily values of headlines did not Granger cause values of interventions in the future (F = .50, *p* = .48); interventions did not Granger cause future daily values of headlines (F = 2.64, *p* = .10).

**C2. NEWS COVERAGE OF SUICIDES AT THE STUDY SITE AND NEARBY CLIFFS WITH METHOD AND/OR LOCATION DETAILS IN THE HEADLINE (N=350)**

***Are media stories of suicides at the study site and nearby cliffs where method and/or location are in the headline associated with suspected suicides at the location and immediately adjacent cliffs?***

Study site news and suspected suicides

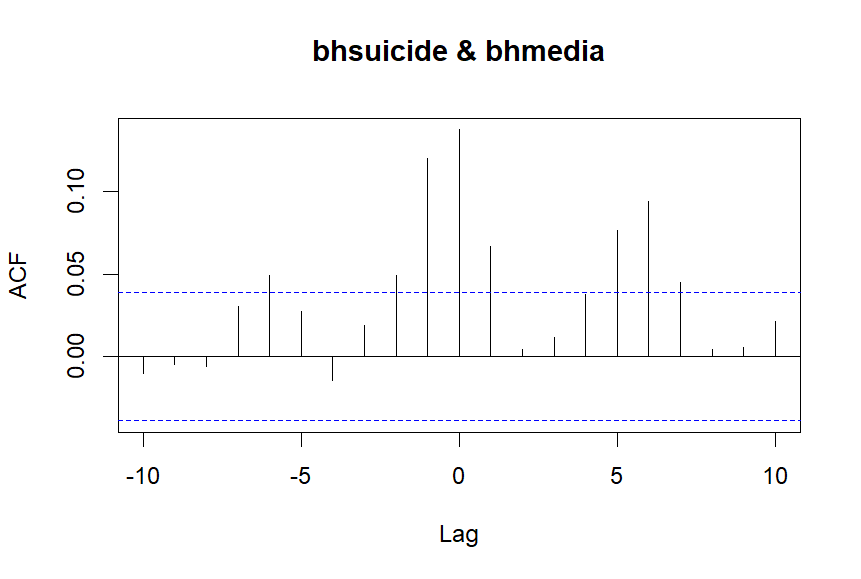

An increase in headlines is associated with an increase in suicides on the day (r = .14), one day (r = .12), two days (r = .05) and six days (r = .05) later. An increase in suicides is associated with an increase in headlines a day later (r = .07) and five to six days later.

Correlations by lag

| **Lag** | -6 | -5 | -4 | -3 | -2 | -1 | 0 | 1 | 2 | 3 | 4 | 5 | 6 |
| --- | --- | --- | --- | --- | --- | --- | --- | --- | --- | --- | --- | --- | --- |
| **Correlation** | .05 | .03 | -.02 | .02 | .05 | .12 | .14 | .07 | .00 | .01 | .04 | .08 | .09 |

Daily values of headlines Granger cause values of suicides in the future (F = 10.58, *p* < .001); suicides Granger cause future daily values of headlines (F = 11.59, *p* < .001).

***Are media stories of suicides at the study site where method and/or location are in the headline associated with crisis interventions at the location?***

Study site news and interventions

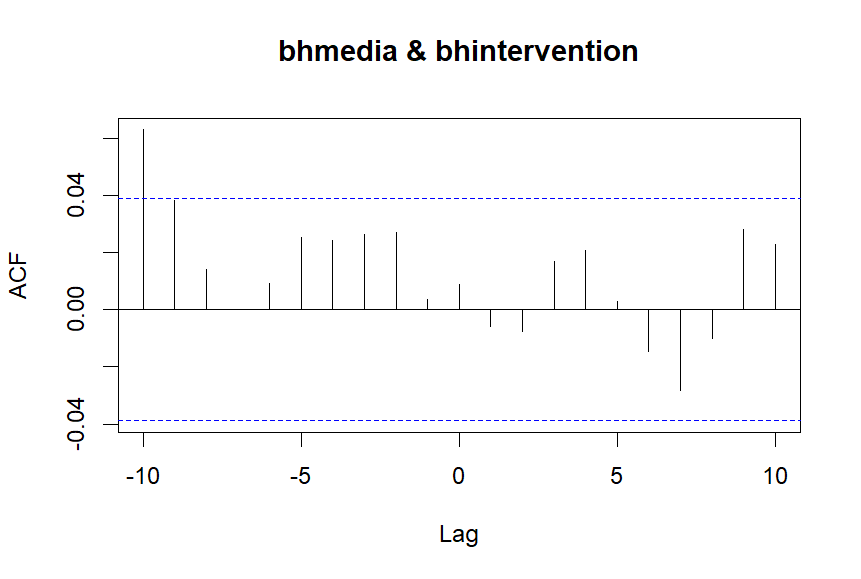

An increase in headlines was not associated with interventions. An increase in interventions was associated with an increase in suicides 10 days later (r = .06).

Correlations by lag

| **Lag** | -10 | -9 | -8 | -7 | -6 | -5 | -4 | -3 | -2 | -1 | 0 | 1 | 2 |
| --- | --- | --- | --- | --- | --- | --- | --- | --- | --- | --- | --- | --- | --- |
| **Correlation** | .06 | .04 | .01 | .0 | .01 | .03 | .02 | .03 | .03 | .00 | .01 | -.01 | -.01 |

Daily values of headlines did not Granger cause values of interventions in the future (F = .32, *p* = .57); interventions did not Granger cause future daily values of headlines (F = .01, *p* = .91).

### D. Associations between news reports of suicidal behaviour at the study site and crisis interventions and suspected suicides at the location and immediately adjacent cliffs, in 2018-19 vs. 2020-23

**D1. ASSOCIATIONS IN 2018-20**

2018 and 2019 were the only years in the broader study period in which a single incident was reported over 10 times. These ‘repeated’ stories alone accounted for a total of 140 reports, i.e. just over a third of all reports of suicides at the study site between 2017 and 2023. They were also the only years for which there are several weeks (six in total) with over 10 stories relating to suicides at this specific coastal location.

**Daily news coverage of suicides at the study site in 2018-19 and suspected suicides at the location and immediately adjacent cliffs**

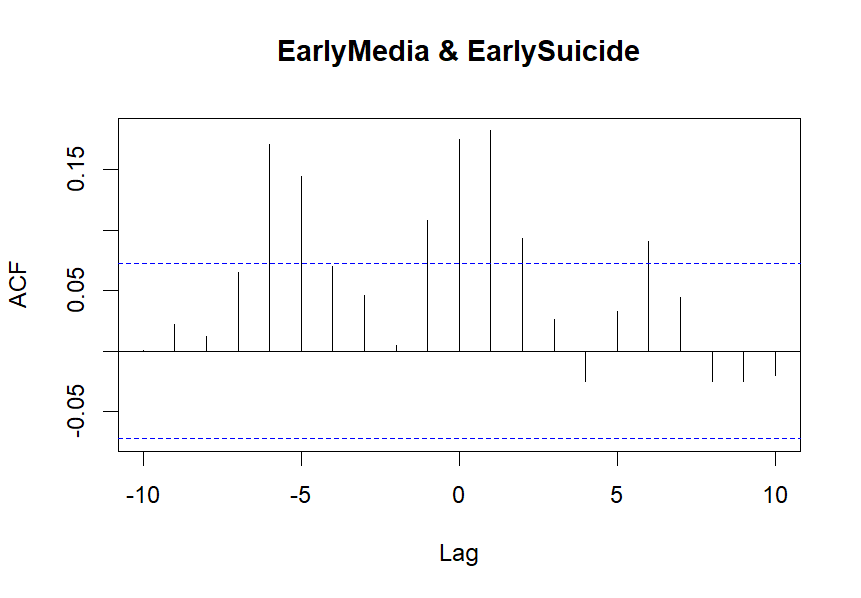

An increase in headlines is associated with an increase in suicides on the day (r = .18), one day (r = .18), two days later (r = .09) and six days later (r = .09). An increase in suicides is associated with an increase in headlines a day later (r = .11) and five to six days later.

Correlations by lag

| Lag | -6 | -5 | -4 | -3 | -2 | -1 | 0 | 1 | 2 | 3 | 4 | 5 | 6 |
| --- | --- | --- | --- | --- | --- | --- | --- | --- | --- | --- | --- | --- | --- |
| Correlation | .17 | .14 | .07 | .05 | .00 | .11 | .18 | .18 | .09 | .03 | -.03 | .03 | .09 |

Daily values of headlines Granger cause values of suicides in the future (F = 8.26, *p* < .01); suicides Granger cause future daily values of headlines (F = 10.46, *p* = .001).

**Daily news coverage of suicides at the study site in 2018-19 and crisis interventions at the location**

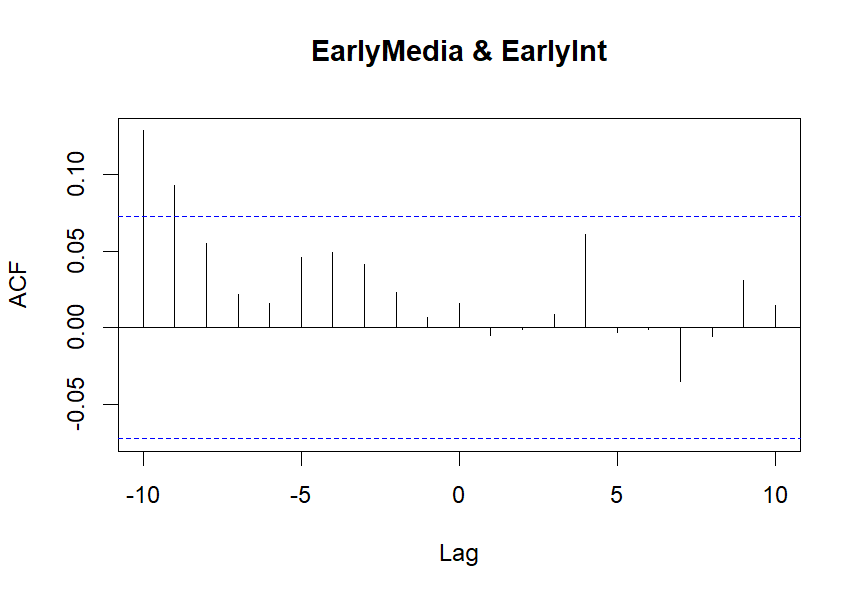

An increase in interventions is associated with an increase in headlines nine days (.09) and ten days later (r = .13). An increase in headlines is not associated with an increase in interventions.

Correlations by lag

| **Lag** | -10 | -9 | -8 | -7 | -6 | -5 | -4 | -3 | -2 | -1 | 0 | 1 | 2 |
| --- | --- | --- | --- | --- | --- | --- | --- | --- | --- | --- | --- | --- | --- |
| **Correlation** | .13 | .09 | .06 | .02 | .02 | .05 | .05 | .04 | .02 | .01 | .02 | -.01 | .00 |

Interventions did not Granger cause future values of headlines (F = .02, *p* = .90); headlines did not Granger cause interventions (F = .19, *p* = .67).

**D2. ASSOCIATIONS IN 2020-23**

Compared to 2018-19, there were fewer reports, fewer stories being covered in the news on multiple occasions, and no headlines making explicit reference to multiple deaths at the site in 2020-23.

**Daily news coverage of suicides at the study site in 2020-23 and suspected suicides at the location and immediately adjacent cliffs**

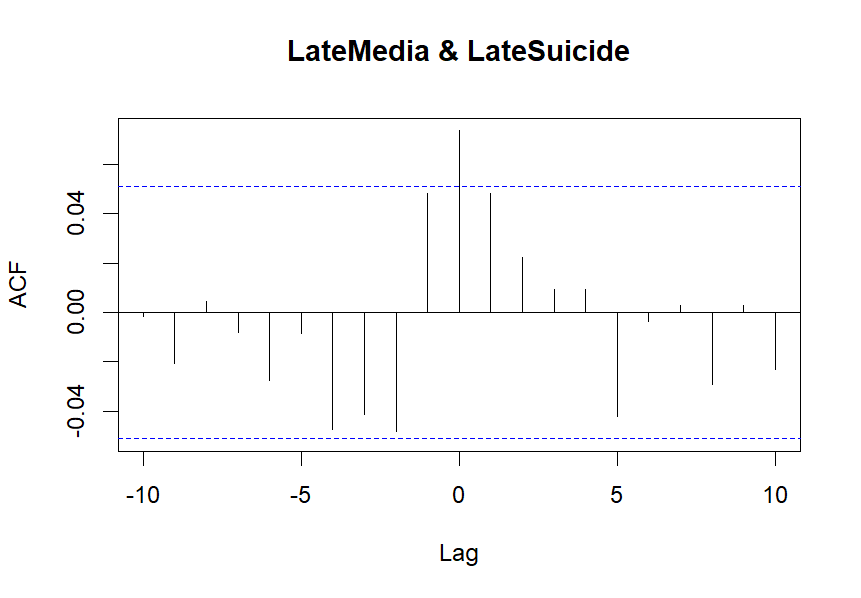

An increase in headlines is associated with an increase in suicides on the day (r = .18).

Correlations by lag

| **Lag** | -7 | -6 | -5 | -4 | -3 | -2 | -1 | 0 | 1 | 2 | 3 | 4 | 5 |
| --- | --- | --- | --- | --- | --- | --- | --- | --- | --- | --- | --- | --- | --- |
| **Correlation** | -.01 | -.03 | -.01 | -.05 | -.04 | -.05 | .05 | .07 | .05 | .02 | .01 | .01 | -.04 |

Daily values of headlines did not Granger cause values of suicides in the future (F = 2.15, *p* = .14); suicides did not Granger cause future daily values of headlines (F = 2.95, *p* = .09).

**Daily news coverage of suicides at the study site in 2020-23 and crisis interventions at the location**

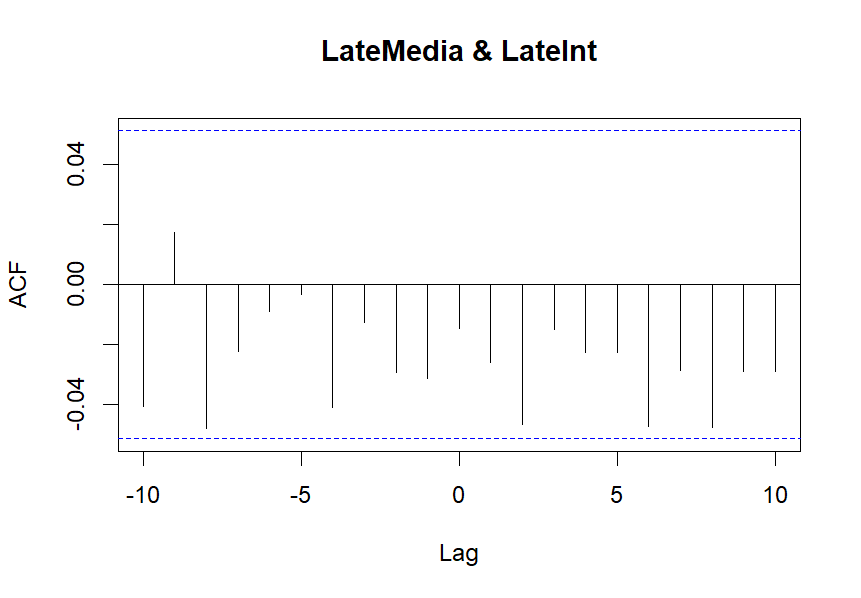

Headlines and interventions were not associated.

Correlations by lag

| **Lag** | -10 | -9 | -8 | -7 | -6 | -5 | -4 | -3 | -2 | -1 | 0 | 1 | 2 |
| --- | --- | --- | --- | --- | --- | --- | --- | --- | --- | --- | --- | --- | --- |
| **Correlation** | -.04 | .02 | -.05 | -.02 | -.01 | .00 | -.04 | -.01 | -.03 | -.03 | -.02 | -.03 | -.05 |

Interventions did not Granger cause future values of headlines (F = 1.26, *p* = .26); headlines did not Granger cause future values of interventions (F = .84, *p* =.36).
